# Causal association of COVID-19 with brain structure changes: Findings from a non-overlapping 2-sample Mendelian randomization study

**DOI:** 10.1101/2023.07.16.23292735

**Authors:** Pingjian Ding, Rong Xu

**Affiliations:** Center for Artificial Intelligence in Drug Discovery, School of Medicine, Case Western Reserve University, Cleveland, Ohio, USA

**Keywords:** COVID-19, Mendelian randomization, brain structure, cortical area, subcortical area

## Abstract

Recent cohort studies suggested that SARS-CoV-2 infection is associated with changes in brain structure. However, the potential causal relationship remains unclear. We performed a two-sample Mendelian randomization analysis to determine whether genetic susceptibility of COVID-19 is causally associated with changes in cortical and subcortical areas of the brain. This 2-sample MR (Mendelian Randomization) study is an instrumental variable analysis of data from the COVID-19 Host Genetics Initiative (HGI) meta-analyses round 5 excluding UK Biobank participants (COVID-19 infection, N=1,348,701; COVID-19 severity, N=1,557,411), the Enhancing NeuroImaging Genetics through Meta Analysis (ENIGMA) Global and regional cortical measures, N=33,709; combined hemispheric subcortical volumes, N=38,851), and UK Biobank (left/right subcortical volumes, N=19,629). A replication analysis was performed on summary statistics from different COVID-19 GWAS study (COVID-19 infection, N=80,932; COVID-19 severity, N=72,733). We found that the genetic susceptibility of COVID-19 was not significantly associated with changes in brain structures, including cortical and subcortical brain structure. Similar results were observed for different (1) MR estimates, (2) COVID-19 GWAS summary statistics, and (3) definitions of COVID-19 infection and severity. This study suggests that the genetic susceptibility of COVID-19 is not causally associated with changes in cortical and subcortical brain structure.

## Introduction

COVID-19 has long-lasting adverse effects on multiple organ systems, including brain [1-3]. A recent cohort study of MRI (magnetic resonance imaging) suggests that SARS-CoV-2 infection was associated with changes in brain structure [2]. However, this study has limitations inherent in observational epidemiological studies including potential reverse causation and confounding bias [4].

Tian et al. [5] reported that the cortical thickness was recovered in COVID-19 cases after a 10-month recovery period, suggesting that the associations between COVID-19 and brain structures are time-dependent. In this study, we employed 2-sample MR (Mendelian randomization) studies to investigate whether genetic susceptibility of COVID-19 is causally associated with changes in cortical and subcortical areas of the brain, including global and 34 regional cortical thickness and surface area as well as 7 combined hemispheric and 17 left/right subcortical volume.

## Methods

This non-overlapping 2-sample MR study was based on GWAS (genome-wide association study) summary statistics of COVID-19 infection and severity (COVID-19 infection, N=1,348,701; COVID-19 severity, N=1,557,411, available from https://www.covid19hg.org/results/r5/) and GWAS summary statistics of brain structure (Global and regional cortical measures [6], N=33,709, available from http://enigma.ini.usc.edu/research/download-enigma-gwas-results ; combined hemispheric subcortical volumes [7], N=38,851, available from http://enigma.ini.usc.edu/research/download-enigma-gwas-results ; left/right subcortical volumes [8], N=19,629, available from https://github.com/BIG-S2/GWAS). The GWAS summary-level data for COVID-19 were obtained from COVID-19 Host Genetics Initiative meta-analyses round 5 (release date: January 18, 2021) [9], which excludes participants from UK Biobank and ensures no overlapping samples between COVID-19 and brain structure GWAS studies. COVID-19 infection was based on testing positive for COVID-19 versus population controls and COVID-19 severity was based on hospitalization of patients with COVID-19 versus population controls [10, 11]. The original GWAS investigations of brain structures were conducted with participants in Enhancing NeuroImaging Genetics through Meta Analysis (ENIGMA) and UK Biobank. The outcome measures included global and 34 regional cortical measures (thickness and surface area) [6] as well as 7 combined hemispheric [7] and 17 left/right subcortical volumes [8] (amygdala, hippocampus, accumbens, putamen, pallidum, thalamus, insula, caudate, and brain stem (combined volumes)). Included GWAS studies have obtained ethical approval from the corresponding ethics review boards.

Instrumental SNPs (single nucleotide polymorphisms) were extracted from GWAS summary statistics of COVID-19 at P<5*10^−6^, ensuring the enough SNPs for estimates. All SNPs were clumped at a window size of 10,000kb and linkage disequilibrium (LD) r2=0.001. We searched appropriate proxies for those SNPs that are not available in the brain structure (r2≥0.8). We harmonized COVID-19 and brain structure data and removed palindromic SNPs with minor allele frequency above 0.42. The Steiger filtering method [12] was implemented to discard SNPs exhibiting reverse causation. The MR-PRESSO method [13] was performed to remove outlier SNPs that may cause horizontal pleiotropy. We conducted inverse-variance weighted (IVW) [14] for the MR main estimate, and MR-Egger [15] and weighted median [16] are used as complementary methods to assess the causality for sensitivity analyses. The beta coefficient and 95% confidence intervals were used to describe the strength of the effect of COVID-19 susceptibility to the changes in brain structures. We performed extensive sensitivity analyses. First, we employed Cochran’s Q statistic and MR-Egger intercept to evaluate heterogeneity among SNPs and pleiotropy, respectively. Second, we conducted the leave-one-out analysis to evaluate single SNP effects.

We validated the findings using summary statistics from AncestryDNA (available from https://rgc-covid19.regeneron.com) [17] with different definitions of COVID infection and severity [10, 11], which included COVID-19 infection, including 1) Covid vs. population; 2) Covid vs. negative; 3) Covid not hospitalized vs. controls, and COVID-19 severity, including 1) hospitalized Covid vs. controls; 2) severity covid vs. controls; 3) hospitalized covid vs. not hospitalized covid.

## Results

We found no significant causal effects of COVID-19 on global cortical thickness (COVID-19 infection: beta 0.003, 95% CI -0.006 to 0.012; COVID-19 severity: beta 0.001, 95% CI -0.001 to 0.004) or on surface area (COVID-19 infection: beta 628.9, 95% CI -731.9 to 1989.7; COVID-19 severity: beta -71.8 95% CI -442.4 to 298.9) (Table 1). Consistently, our study couldn’t find causal associations of COVID-19 infection or severity with changes in 34 regional cortical thickness and surface area (Figure 1). The findings remained null on the combined hemispheric subcortical volumes with genetic susceptibility of COVID-19 infection and severity. Similar null results were obtained for left/right subcortical volumes. Sensitivity analyses suggested that these null results were not due to horizontal pleiotropy (Egger intercept P value > .05) and heterogeneity (Cochran Q P > .05). The null finding was not sensitive to specific SNP selections (leave-one-out analysis, data not shown). These findings were validated in independent GWAS studies of COVID-19 that used different definitions for COVID-19 infection and severity (Figure 2).

**Table.** Results of MR analyses of COVID-19 on global cortical and combined hemispheric subcortical structures.

| Outcome | MR method | COVID-19 infection (#SNPs: 16) |  |  |  | COVID-19 severity (#SNPs:30) |  |  |  |
| --- | --- | --- | --- | --- | --- | --- | --- | --- | --- |
|  |  | MR P | Beta (95% CI) | Q P | Egger P | MR P | Beta (95% CI) | Q P | Egger P |
| Global cortical brain structure |  |  |  |  |  |  |  |  |  |
| thickness | IVW | 0.46 | 0.003(-0.006,0.012) | 0.97 |  | 0.26 | 0.001(-0.001,0.004) | 0.99 |  |
|  | Weighted median | 0.19 | 0.008(-0.004,0.019) |  |  | 0.68 | 0.001(-0.003,0.004) |  |  |
|  | MR Egger | 0.7 | -0.009(-0.052,0.034) | 0.96 | 0.59 | 0.43 | 0.002(-0.003,0.008) | 0.99 | 0.73 |
| Surface area | IVW | 0.37 | 628.9(-731.9,1989.7) | 1 |  | 0.7 | -71.8(-442.4,298.9) | 0.86 |  |
|  | Weighted median | 0.42 | 672.5(-974.2,2319.3) |  |  | 0.86 | -47.8(-582.3,486.8) |  |  |
|  | MR Egger | 0.75 | -994.0(-6878.8,4890.7) | 1 | 0.6 | 0.21 | -555.1(-1392.0,281.9) | 0.9 | 0.22 |
| Combined hemispheric subcortical structures |  |  |  |  |  |  |  |  |  |
| accumbens | IVW | 0.91 | 0.005(-0.077,0.086) | 0.99 |  | 0.48 | 0.01(-0.018,0.039) | 1 |  |
|  | Weighted median | 0.93 | 0.005(-0.098,0.107) |  |  | 0.61 | 0.01(-0.028,0.048) |  |  |
|  | MR Egger | 0.8 | -0.027(-0.227,0.172) | 0.99 | 0.74 | 0.95 | 0.002(-0.059,0.063) | 0.99 | 0.77 |
| amygdala | IVW | 0.61 | 0.021(-0.061,0.103) | 0.96 |  | 0.58 | 0.007(-0.018,0.033) | 0.95 |  |
|  | Weighted median | 0.62 | 0.027(-0.079,0.132) |  |  | 0.17 | 0.024(-0.011,0.06) |  |  |
|  | MR Egger | 0.56 | 0.067(-0.148,0.282) | 0.94 | 0.67 | 0.82 | 0.007(-0.051,0.065) | 0.93 | 0.99 |
| brainstem | IVW | 0.87 | -0.007(-0.094,0.08) | 0.98 |  | 0.74 | 0.006(-0.027,0.038) | 0.97 |  |
|  | Weighted median | 0.51 | -0.038(-0.15,0.075) |  |  | 0.74 | -0.007(-0.05,0.035) |  |  |
|  | MR Egger | 0.79 | -0.033(-0.265,0.198) | 0.97 | 0.82 | 0.48 | -0.026(-0.094,0.043) | 0.98 | 0.33 |
| caudate | IVW | 0.73 | 0.015(-0.07,0.1) | 0.99 |  | 0.86 | -0.002(-0.027,0.023) | 0.9 |  |
|  | Weighted median | 0.74 | 0.018(-0.086,0.121) |  |  | 0.9 | 0.002(-0.033,0.038) |  |  |
|  | MR Egger | 0.93 | -0.01(-0.225,0.205) | 0.97 | 0.81 | 0.92 | 0.003(-0.056,0.062) | 0.86 | 0.85 |
| pallidum | IVW | 0.27 | -0.045(-0.125,0.035) | 0.98 |  | 0.64 | -0.006(-0.032,0.02) | 1 |  |
|  | Weighted median | 0.18 | -0.069(-0.169,0.031) |  |  | 0.33 | -0.017(-0.053,0.018) |  |  |
|  | MR Egger | 0.23 | -0.138(-0.346,0.07) | 1 | 0.37 | 0.33 | -0.029(-0.086,0.028) | 1 | 0.39 |
| putamen | IVW | 0.49 | 0.027(-0.051,0.105) | 0.98 |  | 0.9 | 0.002(-0.025,0.029) | 0.97 |  |
|  | Weighted median | 0.62 | 0.024(-0.073,0.122) |  |  | 0.68 | 0.008(-0.029,0.044) |  |  |
|  | MR Egger | 0.66 | 0.052(-0.165,0.268) | 0.96 | 0.82 | 0.5 | 0.022(-0.039,0.083) | 0.96 | 0.48 |
| thalamus | IVW | 0.76 | 0.011(-0.062,0.085) | 0.99 |  | 0.71 | 0.005(-0.021,0.031) | 1 |  |
|  | Weighted median | 0.51 | 0.031(-0.061,0.123) |  |  | 0.57 | 0.01(-0.024,0.043) |  |  |
|  | MR Egger | 0.72 | 0.037(-0.16,0.235) | 0.98 | 0.79 | 0.79 | 0.008(-0.052,0.068) | 0.99 | 0.9 |

**Figure 1.**
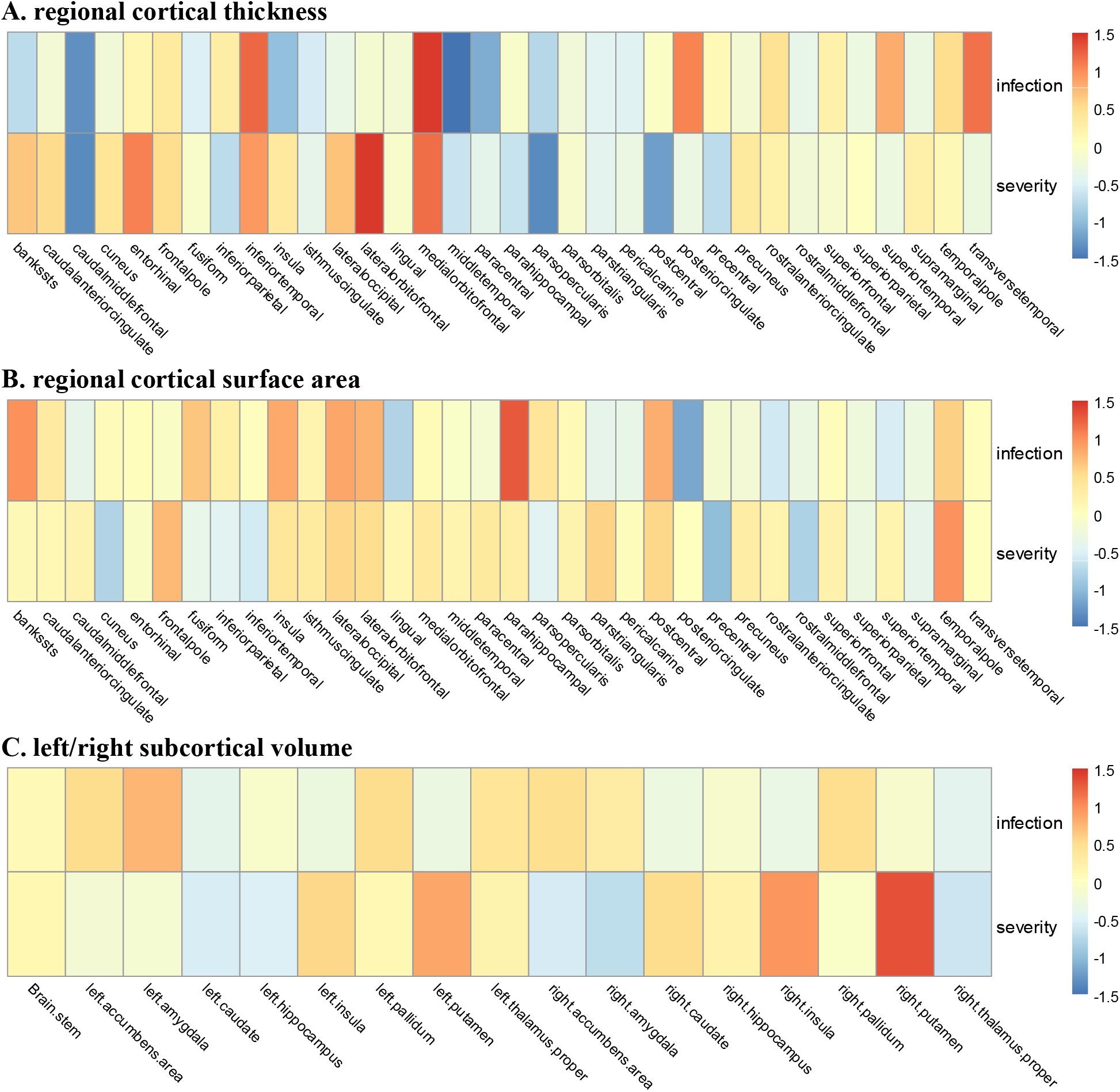
Inverse variance weighted effect estimates (Z Score) for the association between COVID-19 and regional cortical thickness and surface area as well as left/right subcortical volume

**Figure 2.**
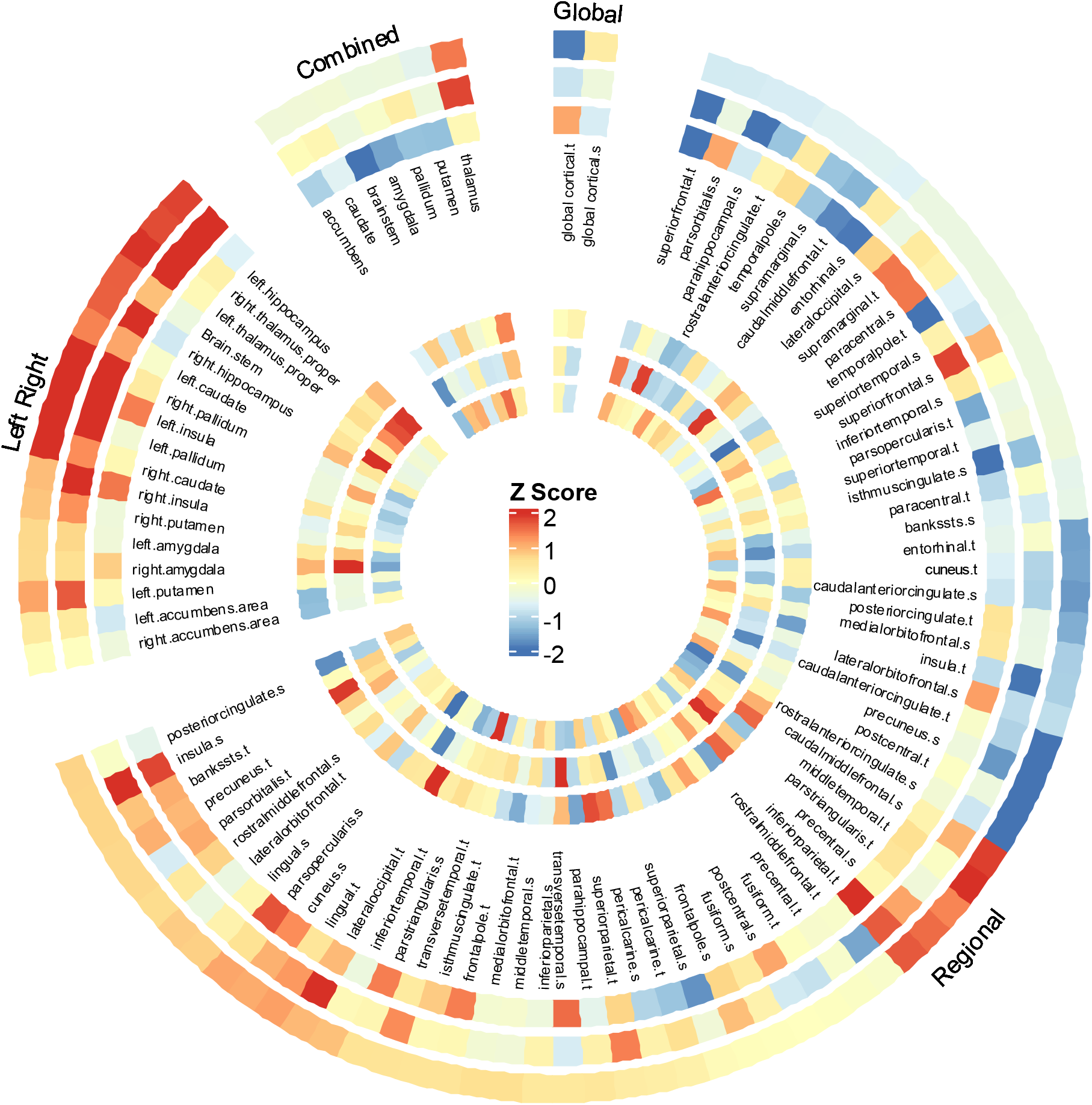
IVW effect estimates (Z Score) using different GWAS summary statistics (COVID-19 GWAS study of AncestryDNA) based on different definitions for COVID-19 infection and severity. The outer ring to the inner ring: 1) Covid vs. population; 2) Covid not hospitalized vs. controls; 3) Covid vs. negative, 4) hospitalized covid vs. not hospitalized covid, 5) hospitalized Covid vs. controls, 6) severity covid vs. controls.

## Discussion

Our non-overlapping 2-sample MR analysis suggests that genetic susceptibility to COVID-19 including both infection and severity, is not causally associated with brain structure changes. Recent cohort studies have shown an association between COVID-19 and subsequent brain structure changes [2, 5]. However, these observed associations are dependent on time elapsed from initial COVID-19 infection and on the SARS-CoV-2 virus strains [4]. Taken together, our MR study and existing cohort studies collectively indicate that the observed brain structures changes following COVID-19 infections are not causal and may have been mediated by modifiable factors other than genetic susceptibility, which are potentially reversible by targeting those modifiable mediators.

Our study has limitations. First, we did not examine the causal effects of genetic susceptibility to COVID-19 on brain structure alterations in specific populations stratified by age, gender, race/ethnicity, virus variants, or comorbidities, due to the lack of such information in the summary statistics data. Second, the GWAS summary statistics from COVID-19 HGI and AncestryDNA were derived from individuals of European ancestry. Further research is warranted to examine the causal associations of COVID-19 with brain structure changes in ethnic and racial minorities in the US who were disproportionately affected by the pandemic [18]. Third, potential selection bias may introduce spurious effect sizes between COVID-19 and brain structure alteration. Finally, considering olfaction and cognitive disorders have improved over time and the cortical thickness was recovered after a 10-month recovery period among COVID-19 patients [5], it is needed to assess the effect of the genetically predicted COVID-19 on brain structure changes at different time points. However, our study did not directly investigate whether the alteration of brain structure in patients with COVID-19 depends on the time period, as we don’t have summary statistics of brain structure at a single time point from COVID-19 infection.

## Author Contribution

Pingjian Ding designed experiments, performed MR analysis, and wrote the manuscript. Rong Xu reviewed and edited the manuscript.

## Acknowledgment

The authors thank the participants from all cohorts who contributed to this study. This work has been supported by the NIH National Institute of Aging (grants nos. AG057557, AG061388, AG062272, AG076649) and the Clinical and Translational Science Collaborative (CTSC) of Cleveland (TR002548).

## Data availability of statement

The summary statistics in this study can be obtained from the original genome-wide association studies. Any other data generated during the analysis can be requested from the authors.

## Ethics approval statement

This study relied on multiple publicly available GWAS summary statistics on COVID-19 and brain structure. Ethical approval was obtained in all original studies.

